# Coronary microvascular resistance comparison of coronary branches with and without additional vascular diameter

**DOI:** 10.1101/2023.02.28.23286601

**Authors:** Takahiro Muroya, Hiroaki Kawano, Fumi Yamamoto, Koji Maemura

**Author notes:** Address for correspondence: Hiroaki Kawano, Department of Cardiovascular Medicine, Nagasaki University Graduate School of Biomedical Sciences, 1-7-1 Sakamoto, Nagasaki 852-8501, Japan. **Conflict of interest:** none.

## Abstract

**Background:** Measurement of coronary microvascular resistance (MR) is essential for diagnosing nonocclusive coronary artery ischemia, but whether coronary branches of different diameters can be similarly assessed using hyperemic microvascular resistance index (hMVRI) calculated from average peak velocity (APV) remains unclear.

**Objectives:** We investigated the relationship between coronary arteries of different diameters and hMVRI.

**Method:** Thirty patients with suspected angina pectoris and nonobstructive coronary stenosis with fractional flow reserve >0.8 underwent evaluation of all coronary arteries using a Doppler velocity and pressure-equipped guidewire. Vessel diameter (D_QCA_) was analyzed by quantitative coronary angiography (QCA). Coronary blood flow (CBF_QCA_) was calculated as πD_QCA_^2^/4 (0.5×APV) and hMVRI_QCA_ as distal coronary pressure (Pd) divided by CBFD_QCA_ during maximal hyperemia.

**Results:** The hMVRI was significantly higher for the right coronary artery (RCA) than for the left anterior descending artery (LAD), but no significant differences between arteries were seen for CBF_QCA_ and hMVRI_QCA_. Although the correlation between CBF_QCA_ and APV was weak in all arteries, CBF_QCA_ divided into three groups according to D_QCA_ showed very strong correlations with APV. Slopes of the straight line between APV and CBF_QCA_ for small-, middle-, and large-diameter groups were 0.48, 0.30, and 0,21, respectively, with slope decreasing as diameter increased. The correlation between APV and CBF_QCA_ was high for LAD and RCA, but weak for the left circumflex artery.

**Conclusions:** Evaluation of MR in coronary branches requires consideration of vessel diameter.

## Clinical Perspective

### What is new?

• Measurement of coronary microvascular resistance (MR) is essential for diagnosing nonocclusive coronary artery ischemia, but whether coronary branches of different diameters can be similarly assessed using hyperemic microvascular resistance index (hMVRI) calculated from average peak velocity (APV) remains unclear.
• We investigated the relationship between coronary arteries of different diameters and hMVRI.

### What are the clinical implications?

• The hMVRI was significantly higher for the right coronary artery (RCA) than for the left anterior descending artery (LAD), but no significant differences between arteries were seen for CBF_QCA_ and hMVRI_QCA_.
• Evaluation of MR in coronary branches requires consideration of vessel diameter.

## INTRODUCTION

Although angina pectoris affects approximately 112 million people globally, up to 70% of patients undergoing invasive angiography do not have obstructive coronary artery disease and the main cause is ischemia with no obstructive coronary arteries (INOCA), which can result from heterogeneous mechanisms, including coronary vasospasm and microvascular dysfunction, and is not a benign condition.^1^ Since no currently available modalities allow direct visualization of the human coronary microcirculation in vivo, microvascular evaluation relies on the measurement of parameters that reflect the functional status, such as coronary blood flow (CBF), coronary flow reserve (CFR), and microvascular resistance, using a diagnostic guidewire. Both microvascular and epicardial vasospastic angina are then assessed with the acetylcholine test. Lifestyle modification, risk factor management, and anti-anginal medications have been proposed for the management of INOCA. However, studies of therapies to improve coronary microvascular dysfunction (CMD) have been small and heterogeneous in both design and methodology, and no evidence-based treatments for CMD are currently available. An updated standardization of criteria for microvascular angina (MVA) in patients presenting with angina pectoris or ischemia-like symptoms in the absence of flow-limiting coronary artery disease has been proposed by the COVADIS group and abnormal coronary microvascular resistance indices are among the leading diagnostic parameters. Although these parameters can be investigated using the index of microcirculatory resistance (IMR) or hyperemic microvascular resistance index (hMVRI), evaluation in the catheterization laboratory has become easier in recent years and guidelines recommend IMR and HMR measurements to investigate INOCA. Although many reports^1-20^ have already examined the usefulness and pitfalls of guidewires in the examination of IMR or hMVRI, which coronary artery should be targeted for evaluation in INOCA remains a vexing question. The LAD is the preferred target vessel in many studies, reflecting its myocardial mass and coronary arterial predominance, and much of the evidence for the assessment of physiology in INOCA has been obtained from investigations using the LAD.^7-9^ On the flip side, when the LAD cannot be used for technical reasons, use of the left circumflex artery (LCX) is recommended, followed by the right coronary artery (RCA). The reason for this may be that previous reports have shown that HMR in the RCA is significantly higher than in other coronary branches^14,15^. Average peak velocity (APV) is used instead of coronary blood flow (CBF) for coronary microvascular resistance measurement, because IMR and HMR are used for simple measurements in the cardiac catheterization laboratory. However, whether coronary branches of widely varying diameters can be evaluated equivalently by APV in humans remains highly questionable. The present study examined cases in which all three coronary branches were measured in the same patient to determine differences in coronary microcirculatory resistance in human coronary arteries of different diameters.

## METHODS

### Patient population

The patient population is presented in Figure 1. We evaluated patients with clinical suspicion of stable angina pectoris (SAP) based on the presence of chest pain or chest discomfort. Evaluations were performed using elective coronary angiography and physiological assessments between September 2010 and May 2016. A total of 203 patients with fractional flow reserve (FFR) >0.8 in at least one coronary branch were evaluated. Patients with hypertrophic cardiomyopathy (n=6), moderate-to-severe aortic valve stenosis disease (n=3), coronary artery bypass graft surgery (n=1), atrial fibrillation (n=6), cardiac pacemaker insertion (n=5), recent myocardial infarction (<6 weeks before screening) (n=13), visible collateral development to the perfusion territory of interest (n=3) or left ventricular ejection fraction <50% (n=6) were excluded. As a result, we assessed 281 coronary arteries in 160 patients (Group A), comprising 121 LADs, 83 LCXs, and 77 RCAs. In all 30 of these patients (Group B) in whom FFR was >0.8 in all three coronary branches, coronary microvascular resistance between coronary branches was examined separately using both conventional indices and indices that considered vessel size using quantitative coronary angiography (QCA). All patients provided written informed consent for participation and the Ethics Committee at Ureshino Medical Center approved the protocol for the present study (approval no. 10-10), which was performed in accordance with the principles laid out in the Declaration of Helsinki.

**Figure 1.**
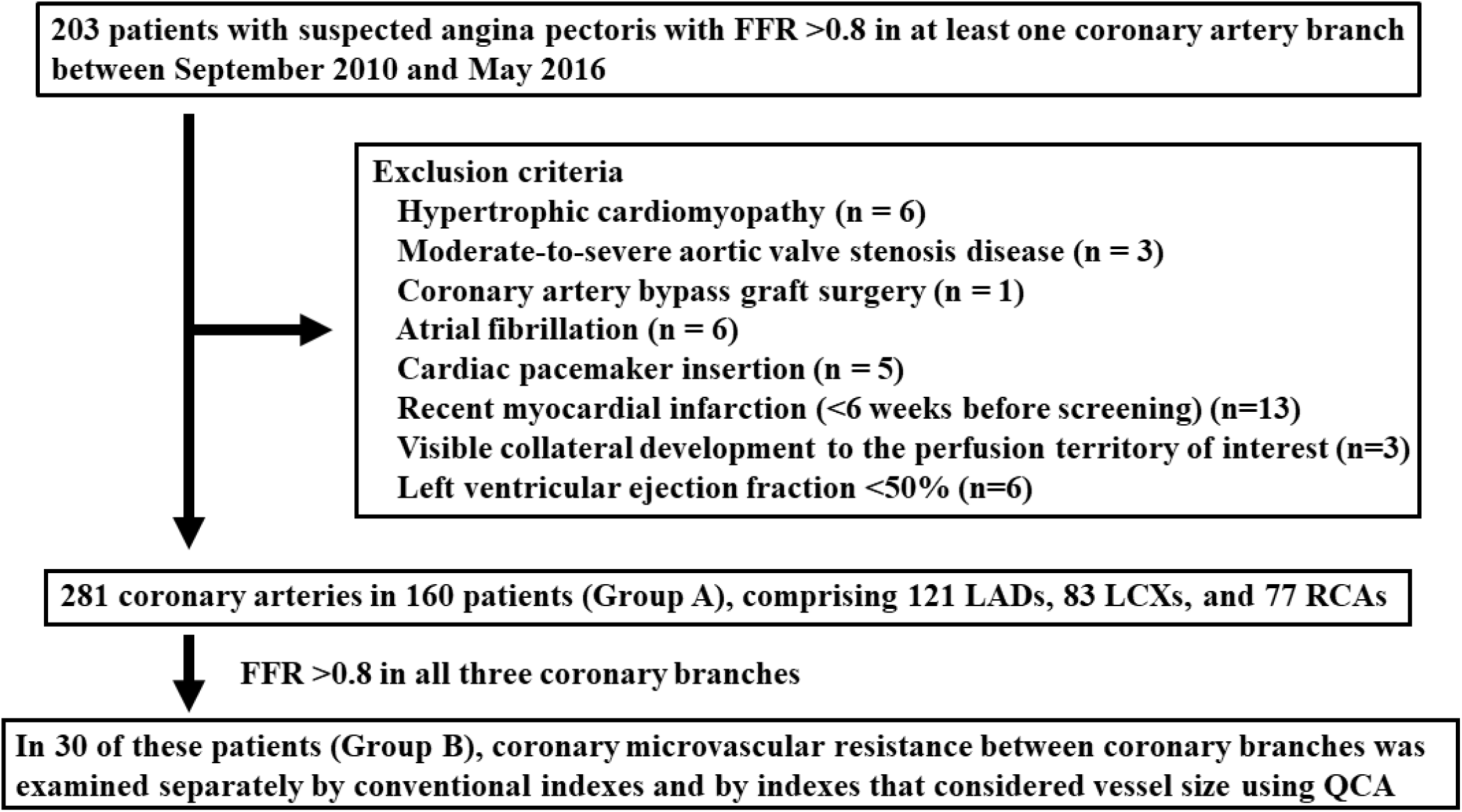
Patient population. We investigated 203 patients with suspected angina pectoris with FFR >0.8 in at least one coronary artery branch between September 2010 and May 2016. Patients with hypertrophic cardiomyopathy (n=6), moderate-to-severe aortic valve stenosis disease (n=3), coronary artery bypass graft surgery (n=1), atrial fibrillation (n=6), cardiac pacemaker insertion (n=5), recent myocardial infarction (<6 weeks before screening) (n=13), visible collateral development to the perfusion territory of interest (n=3) or left ventricular ejection fraction <50% (n=6) were excluded. As a result, we assessed 281 coronary arteries in 160 patients (Group A), comprising 121 LADs, 83 LCXs, and 77 RCAs. In all 30 of these patients (Group B) with FFR >0.8 in all three coronary branches, microvascular resistance between coronary branches was examined separately by conventional indices and by indices that considered vessel size using quantitative coronary angiography (QCA).

### Cardiac catheterization procedure

After coronary angiography, aortic pressure was measured via a 5- or 6-Fr guiding catheter placed in the coronary ostium via a radial or femoral approach. Intracoronary pressure and coronary flow velocity were measured with a 0.014" Doppler velocity and pressure-equipped guidewire (Volcano Corp, San Diego, CA), then the sensor was placed at a site more distal than the distal half of the coronary artery. Hyperemia was induced by papaverine hydrochloride delivered into the coronary artery (12 mg into the left coronary artery and 8 mg into the RCA within 15 s), then blood pressure was recorded at 20 s after the end of administration. FFR was defined as the ratio of mean distal coronary pressure (Pd) to mean aortic pressure (Pa) in the target vessel beyond the lesion during maximal hyperemia.

Hyperemic microvascular resistance index (hMVRI) was calculated as Pd divided by distal APV during maximal hyperemia. In the present study, those coronary arteries showing FFR >0.8 were defined as non-obstructive coronary arteries unaffected by collateral effects^5^, and were investigated. Vessel diameter analyzed by quantitative coronary angiography (D_QCA_) at the measurement site of the wire where blood flow was measured (Figure 2). QCA was performed using a validated densitometric analysis system (Pie Medical Imaging B.V, Maastricht, the Netherlands). Coronary blood flow (CBF_QCA_) was calculated as πD_QCA_^2^/4 (0.5×APV) (D_QCA_, vessel diameter), then hMVRI_QCA_ was calculated as Pd divided by CBF_QCA_ during maximal hyperemia.

**Figure 2.**
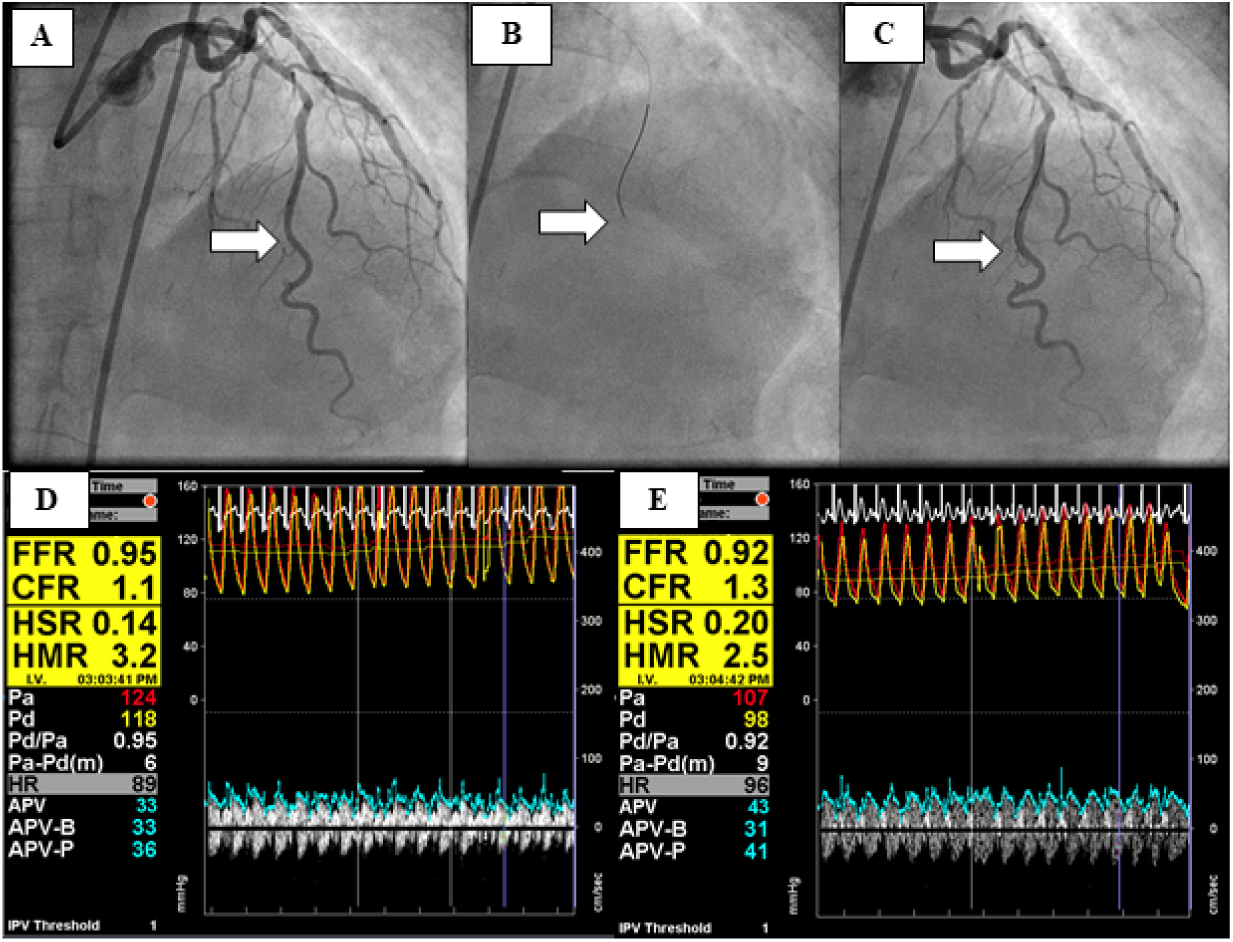
Calculation of coronary physiological values. Hyperemic microvascular resistance index (hMVRI) was calculated as Pd divided by distal APV at maximal hyperemia. Vessel diameter was analyzed by quantitative coronary angiography (QCA) at the site of blood flow measurement by wire (white arrow). QCA was performed with a validated densitometric analysis system (Pie Medical Imaging B.V, Maastricht, the Netherlands). Coronary blood flow (CBF) was calculated as πD^2^/4 (0.5×APV) (D, vessel diameter; APV, average peak velocity). CBF measured by QCA is described as CBF_QCA_, then hMVRI_QCA_ was calculated as Pd divided by CBF_QCA_ during maximal hyperemia. As CBF_QCA_ will be overestimated, hMVRI_QCA_ will be higher than the original HMR. A) Coronary angiography. B) Insertion of the Doppler flow guide wire into the left anterior descending artery. C) Final position of the Doppler flow guide wire as confirmed by contrast injection. D) Physiological measurements at rest. E) Physiological measurements under hyperemic conditions.

In these patients, coronary microvascular dysfunction (CMD) was evaluated using modified criteria from previous reports^2-13^ for the diagnosis of microvascular angina.

Coronary physiological values were calculated as follows:

FFR = Pd / Pa during hyperemia

hMVRI (mmHg/cm/s) = Pd / APV during hyperemia

CBF_QCA_ (ml/min) = πD_QCA_^2^/4 (0.5×APV) (D_QCA_, vessel diameter using QCA)

hMVRI_QCA_ (mmHg/cm/s) = Pd / CBF_QCA_ during hyperemia

### Statistical analysis

All data are expressed as median ± standard deviation or the number and percentage of patients. Non-parametric paired Wilcoxon analysis testing was used to test for associations of intra-coronary physiological values, vessel diameter and CBF_QCA_ between LAD, LCX, and RCA. Associations between APV and CBF_QCA_ in Group B, a small vessel diameter group, middle diameter group, large diameter group, LAD, LCX, and RCA were evaluated using univariate linear regression analysis. Values of *p* < 0.05 were considered statistically significant. Data were statistically analyzed using JMP version 10 software (SAS Institute Inc., Cary, NC, USA).

## RESULTS

### Patient characteristics

Patient characteristics in Groups A and B are presented in Table 1. Median age in Groups A and B was 69.0 years and 74 years, respectively. Men comprised 113 of 160 patients (70.6%) in Group A and 23 of 30 patients (76.7%) in Group B. No significant differences in patient characteristics were seen between Groups A and B, except for in HbA1c. HbA1c was significantly lower in Group B than in Group A (*p*=0.040), but no significant difference in the incidence of diabetes mellitus was identified (*p*=0.301).

**Table 1.** Patient characteristics in Groups A and B.

| Variable | Group A (n=160) | Group B (n=30) | p value |
| --- | --- | --- | --- |
| **General risk factors** |  |  |  |
| Age, y | 69.0±9.6 | 74.0±10.3 | 0.272 |
| Male sex, n (%) | 113 (70.6) | 23 (76.7) | 0.501 |
| Body mass index, kg/m<sup>2</sup> | 24.5±3.9 | 24.5±3.9 | 0.428 |
| **Cardiovascular risk factors** |  |  |  |
| Hypertension, n (%) | 112 (70) | 18 (60.0) | 0.280 |
| Dyslipidemia, n (%) | 110 (68.8) | 17 (56.7) | 0.197 |
| Diabetes mellitus, n (%) | 64 (40) | 9 (30.0) | 0.301 |
| Current smoker, n (%) | 43 (26.9) | 4 (13.3) | 0.115 |
| **Laboratory findings** |  |  |  |
| HDL, mg/dl | 53±16 | 52±15 | 0.900 |
| LDL, mg/dl | 101±31 | 97±29 | 0.296 |
| TG, mg/dl | 113±93 | 97.5±93 | 0.443 |
| Cr, mg/dl | 0.79±0.78 | 0.81±0.84 | 0.471 |
| HbA1c, % | 5.7±1.1 | 5.4±1.0 | 0.040 |
| **CAD** |  |  |  |
| Previous MI, n (%) | 34 (21.3) | 6 (20.0) | 0.878 |
| Previous PCI, n (%) | 85 (53.1) | 20 (66.7) | 0.171 |
| **Medications** |  |  |  |
| β-blocker, n (%) | 21 (13.1) | 4 (13.3) | 0.975 |
| Calcium channel blocker, n (%) | 84 (52.5) | 13 (43.3) | 0.357 |
| ARB or ACE inhibitor, n (%) | 77 (48.1) | 11 (36.7) | 0.248 |
| Statin, n (%) | 98 (61.3) | 17 (56.7) | 0.637 |
| Diuretic, n (%) | 7 (4.4) | 1 (3.3) | 0.794 |
| **UCG findings** |  |  |  |
| LVDd, mm | 47.2±3.7 | 46.9±3.1 | 0.739 |
| LVDs, mm | 29.8±3.8 | 29.0±3.4 | 0.373 |
| LVEF, % | 66.9±6.5 | 67.7±7.1 | 0.301 |

*Note:* Values are presented as n (%) or median ± standard deviation.
Abbreviations: HDL, high-density lipoprotein cholesterol; LDL, low-density lipoprotein cholesterol; TG, triglycerides; Cr, creatinine; HbA1c, hemoglobin A1c; MI, myocardial infarction; PCI, percutaneous coronary intervention; ARB, angiotensin receptor blocker; ACE, angiotensin-converting enzyme inhibitor; UCG, ultrasonic echocardiography; LVDd, left ventricular

### Relationship between coronary physiological assessments by Doppler and pressure sensor-equipped guide-wire and the three coronary arteries

Coronary physiological assessments of Groups A and B are presented in Tables 2 and 3 and Figures 3 and 4 respectively. Median values of FFR, hMVRI, APV, and Pd in Group A were 0.92, 1.9 mmHg/cm/s, 37, and 72 mmHg/cm/s, respectively. Median values of FFR, hMVRI, APV and Pd in Group B were 0.94, 1.9 mmHg/cm/s, 29, and 74 mmHg/cm/s, respectively.

**Figure 3.**
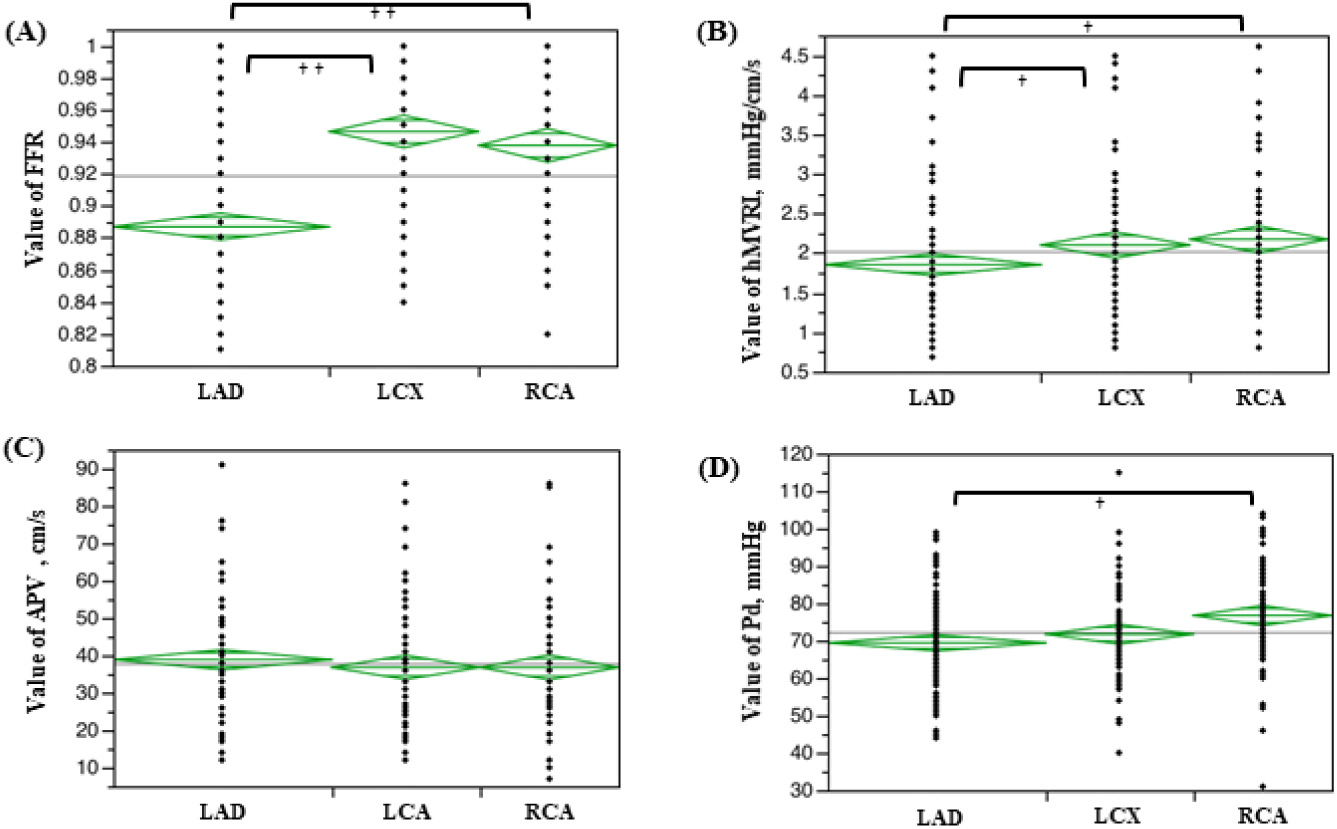
Evaluation of FFR, hMVRI, APV and Pd in Group A. In comparisons between LAD, LCX, and RCA, significant differences in FFR, hMVRI, and Pd are seen in Group A. A) FFR is significantly lower in the LAD than in the LCX or RCA (*p*<0.001). B) hMVRI is significantly lower in the LAD than in the LCX or RCA (*p*=0.003). C) No significant differences in APV are seen between the three coronary branches. D) Pd is significantly lower in the LAD than in the RCA (*p*<0.001). ^†^ *p*<0.050; ^††^ *p*<0.001

**Figure 4.**
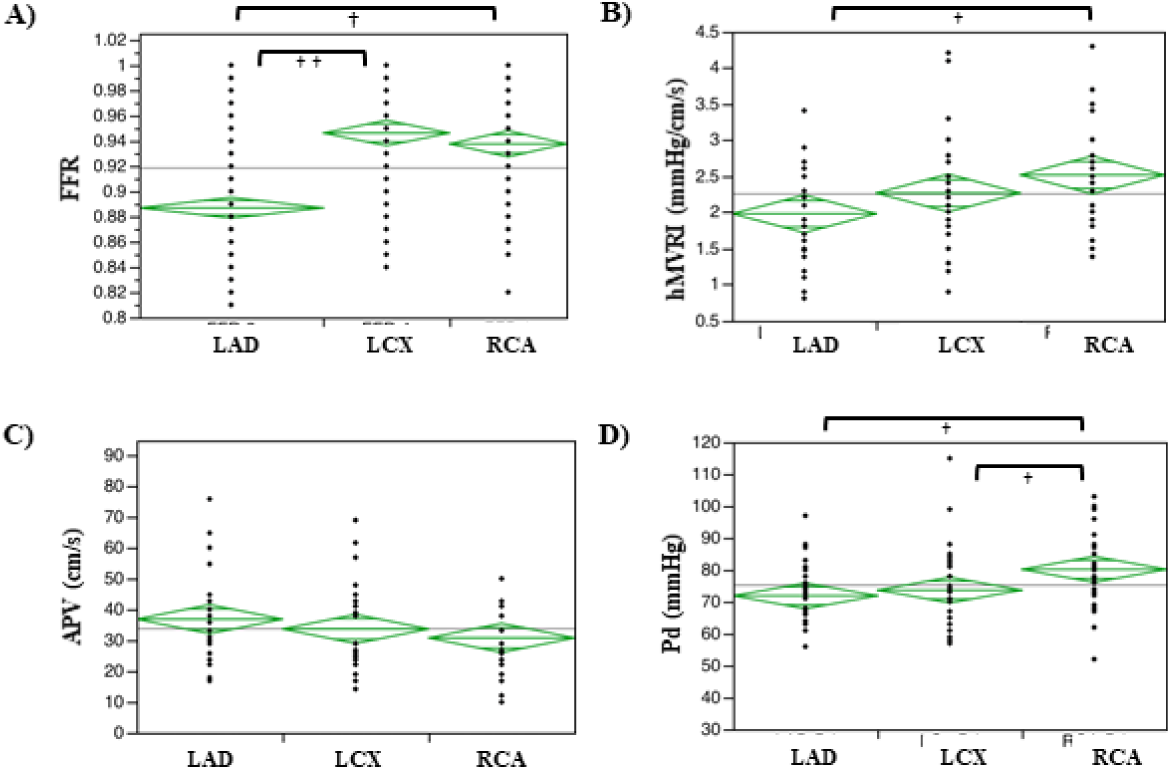
Evaluation of FFR, hMVRI, APV, and Pd in Group B. In comparisons between LAD, LCX, and RCA, significant differences are seen in FFR, hMVRI and Pd in Group B. A) FFR is significantly lower in the LAD than in the LCX or RCA (*p*<0.001). B) hMVRI is significantly lower in the LAD than in the RCA (*p*=0.019). C) No significant differences in APV is seen between the three coronary branches. D) Pd is significantly lower in the LAD and LCX than in the RCA (*p*=0.004). ^†^ *p*<0.05; ^††^ *p*<0.001

**Table 2.** Coronary physiological assessments by Doppler and pressure sensor-equipped guide-wire in Group A.

| Variable | Group A (n=160) | LAD (n=121) | LCX (n=83) | RCA (n=77) | P value |
| --- | --- | --- | --- | --- | --- |
| FFR | 0.92±0.05 | 0.88±0.05 | 0.96±0.05 | 0.94±0.05 | <0.0001 |
| hMVRI, mmHg/cm/s | 1.9±0.8 | 1.7±0.7 | 2.0±0.8 | 2.0±0.8 | 0.003 |
| APV, cm/s | 37±15 | 38±15 | 36±15 | 36±15 | 0.381 |
| Pd, mmHg | 72±12 | 70±12 | 71±12 | 77±13 | <0.0001 |

*Note:* Values are presented as n (%) or median ± standard deviation.
Abbreviations: FFR, fractional flow reserve; hMVRI, hyperemic microvascular resistance index; APV, average peak velocity; Pd, mean distal coronary pressure; LAD, left anterior descending coronary artery; LCX, left circumflex; RCA, right coronary artery.

In terms of the relationships between LAD, LCX, and RCA, similarly significant differences in FFR, hMVRI and Pd were seen in Groups A and B (Tables 2, 3; Figs. 3, 4). The hMVRI was significantly higher for the RCA than for the LAD in both Groups A and B.

**Table 3.**
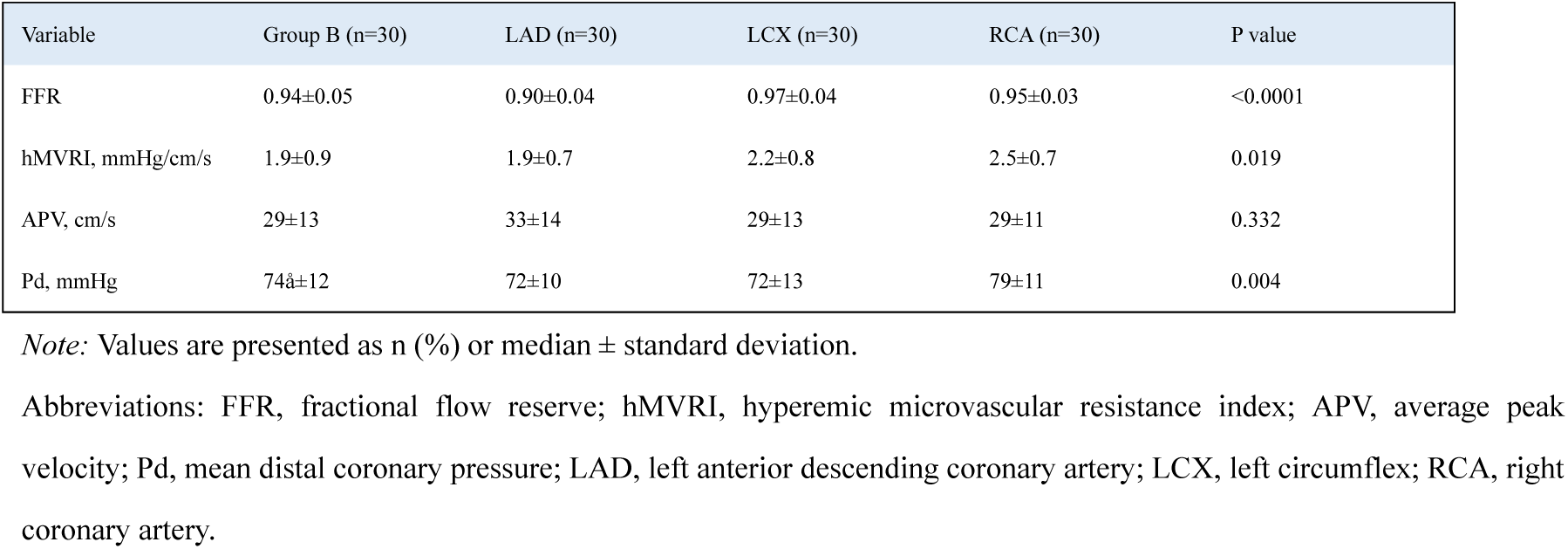
Coronary physiological assessments by Doppler and pressure sensor-equipped guide-wire in Group B.

| Variable | Group B (n=30) | LAD (n=30) | LCX (n=30) | RCA (n=30) | P value |
| --- | --- | --- | --- | --- | --- |
| FFR | 0.94±0.05 | 0.90±0.04 | 0.97±0.04 | 0.95±0.03 | <0.0001 |
| hMVRI, mmHg/cm/s | 1.9±0.9 | 1.9±0.7 | 2.2±0.8 | 2.5±0.7 | 0.019 |
| APV, cm/s | 29±13 | 33±14 | 29±13 | 29±11 | 0.332 |
| Pd, mmHg | 74±12 | 72±10 | 72±13 | 79±11 | 0.004 |

*Note:* Values are presented as n (%) or median ± standard deviation.
Abbreviations: FFR, fractional flow reserve; hMVRI, hyperemic microvascular resistance index; APV, average peak velocity; Pd, mean distal coronary pressure; LAD, left anterior descending coronary artery; LCX, left circumflex; RCA, right coronary artery.

### Relationships between D_QCA_, CBF_QCA_, and hMVRI_QCA_ of LAD, LCX, and RCA in Group B

D_QCA_ was analyzed by QCA at the site of blood flow measurement by diagnostic wire in Group B, with hMVRI re-evaluated as hMVRI_QCA_ and CBF as CBF_QCA_. D_QCA_ of the LAD, LCX, and RCA was 2.27 mm, 2.81 mm, and 2.91 mm, respectively; D_QCA_ was significantly larger for the LCX and RCA than for the LAD (LCX, *p*=0.029; RCA, *p*<0.001), but no significant difference was seen between D_QCA_ of the LCX and RCA (*p*=0.152) (Table 4, Fig. 5A). Unlike the results examined in the APV, no significant differences between LAD, LCX or RCA were seen for CBF_QCA_ or hMVRI_QCA_ (Table 4; Fig. 5B, 5C).

**Figure 5.**
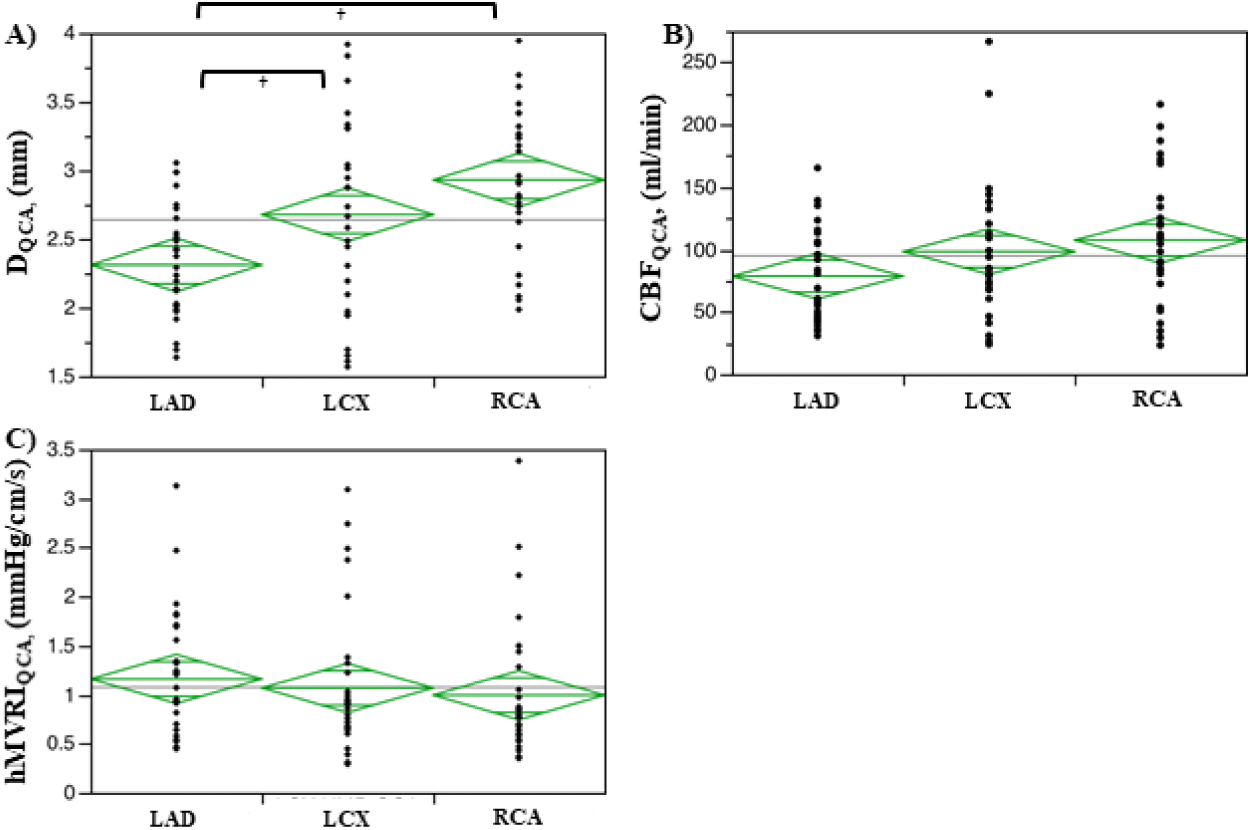
CBF_QCA_ and HMR_QCA_ corrected for vessel diameter by QCA. A) D_QCA_ in the LAD, LCX, and RCA are 2.27 mm. 2.81 mm, and 2.91 mm, respectively. D_QCA_ is significantly larger in the LCX and RCA than in the LAD (LCX, *p*=0.029; RCA,*p*<0.001), but no significant difference is seen between the LCX and RCA (*p*=0.152). B, C) No significant differences in CBF_QCA_ or hMVRI_QCA_ are seen between LAD, LCX, and RCA. ^†^ *p*<0.05; ^††^ *p*<0.001

**Table 4.** Re-evaluation of Group B using HMR_QCA_ and CBF_QCA_.

| Variable | LAD | LCX | RCA | P value |
| --- | --- | --- | --- | --- |
| **D<sub>QCA</sub>, mm** | 2.27±0.39 | 2.81±0.67 | 2.91±0.51 | 0.0002 |
| **CBF<sub>QCA</sub>, ml/min** | 65.0±37.7 | 82.4±59.1 | 106.7±50.9 | 0.100 |
| **hMVRI<sub>QCA</sub>, mmHg/cm/s** | 1.04±0.69 | 0.87±0.74 | 0.78±0.69 | 0.384 |

*Note:* Values are presented as n (%) or median ± standard deviation.
Abbreviations: D, diameter; hMVRI, hyperemic microvascular resistance index; CBF, coronary blood flow; LAD, left anterior descending coronary artery; LCX, left circumflex; RCA, right coronary artery.

### APV and CBF_QCA_ for small D_QCA_, middle D_QCA_, and large D_QCA_ in Group B

We examined the relationship between CBF_QCA_ and APV in Group B, but the correlation was weak (*p*<0.001, R^2^=0.34) (Fig. 6A). We then divided vessels into three equal groups according to D_QCA_: small diameter group, 1.57–2.29 mm; middle diameter group, 2.31–2.9 mm; and large diameter group, 2.92–3.94 mm. We assessed associations between APV and CBF_QCA_ for each group, revealing that CBF_QCA_ in the three groups correlated strongly with APV (small diameter group: R^2^=0.85, *p*<0.001; middle diameter group: R^2^=0.87, *p*<0.001; large diameter group: R^2^=0.80, *p*<0.001) (Fig. 6B–D). The slope of the straight line between APV and CBF_QCA_ for small, middle, and large diameter groups was 0.48, 0.30, and 0.21, respectively, with slope decreasing as diameter increased.

**Figure 6.**
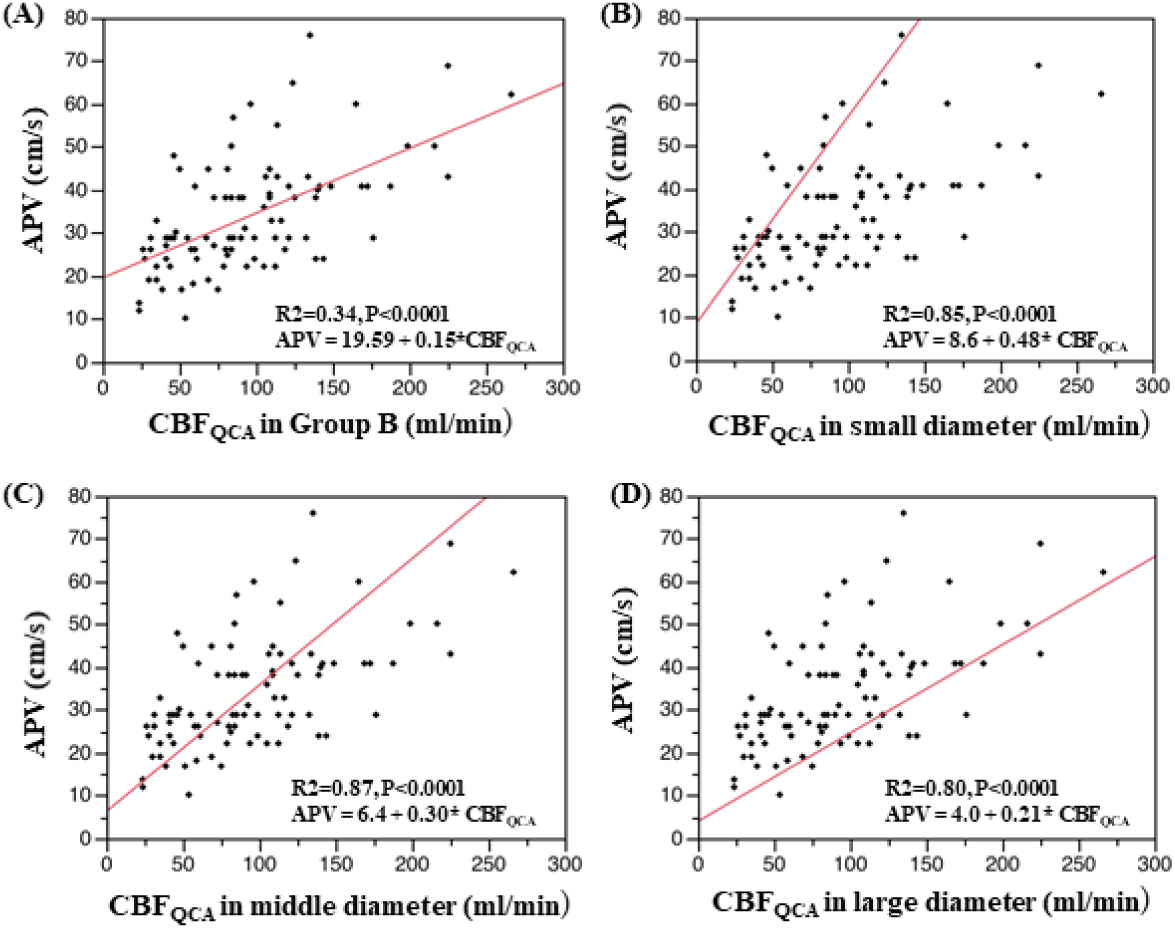
Correlation between APV and CBF_QCA_ in Group B vessels. A) CBF_QCA_ and APV in Group B show a weak correlation (*p*<0.001, R^2^=0.34; APV = 19.59 + 0.15*CBF_QCA_). We divided Group B into three equal groups according to the D_QCA_: small diameter group, 1.57–2.29 mm; middle diameter group, 2.31–2.9 mm; and large diameter group, 2.92–3.94 mm. B–D) The relationships between CBF_QCA_ and APV in the small, middle and large diameter groups are strong (small diameter group: R^2^=0.85, *p*<0.001, APV = 8.6 + 0.48×CBF_QCA_; middle diameter group: R^2^=0.87, *p*<0.001, APV = 6.4 + 0.30×CBF_QCA_; large diameter group: R^2^=0.80, *p*<0.001, APV = 4.0 + 0.21×CBF_QCA_). The slope of the straight line between APV and CBF_QCA_ for small, middle, and large diameters is 0.48, 0.30, and 0,21, respectively, decreasing as diameter increases.

### Correlations between APV and CBF_QCA_ of LAD, LCX, and RCA in Group B

We assessed associations between APV and CBF_QCA_ for each coronary artery branch. The correlation was strong in LAD and RCA (LAD: R^2^=0.52, *p*<0.001; RCA: R^2^=0.57, *p*<0.001) (Fig. 7A, 7C) but the correlation in LCX was weak (LCX: R^2^=0.36, *p*<0.001) (Fig. 7B).

**Figure 7.**
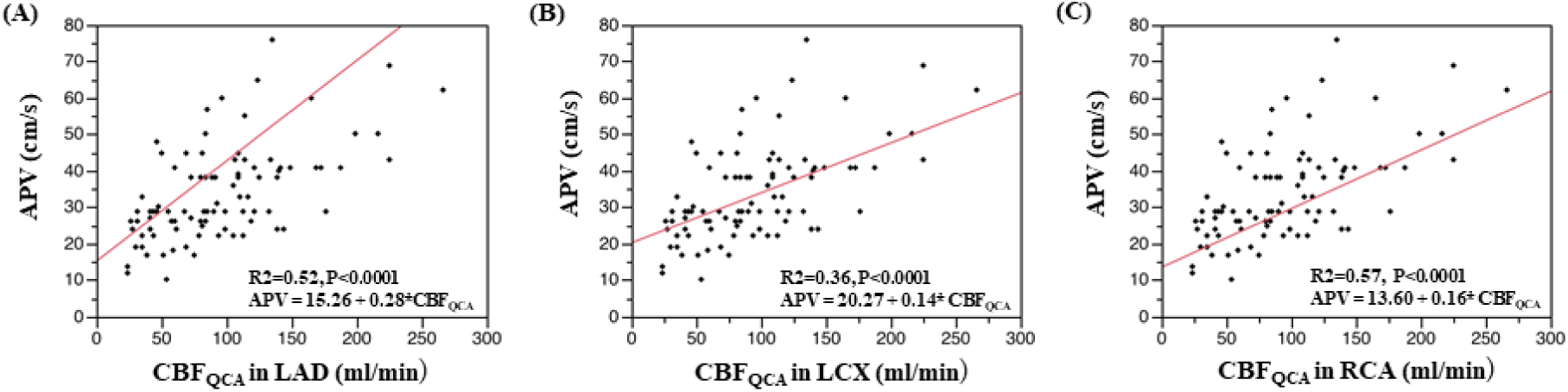
Correlation between APV and CBF_QCA_ in Group B coronary branches. We assessed associations between APV and CBF_QCA_ in vessel branches. We assessed associations between APV and CBF_QCA_ in vessel branches. A, C) Correlations in LAD and RCA are strong (LAD: R^2^=0.52, *p*<0.001, APV = 15.26 + 0.28×CBF_QCA_; RCA: R^2^=0.57, *p*<0.001, APV = 20.27 + 0.14×CBF_QCA_). B) The correlation in LCX is weak (LCX: R^2^=0.36, *p*<0.001, APV = 13.60 + 0.16×CBF_QCA_)

## DISCUSSION

The present study examined whether differences exist in microcirculatory resistance in human coronary arteries of different sizes. The main findings were as follows. First, as reported previously, hMVRI was significantly higher for the RCA than for the LAD in Groups A and B. Second, unlike the results for the APV, no significant differences in hMVRI_QCA_ calculated using D_QCA_ were seen between LAD, LCX, and RCA. Moreover, the correlation between CBF_QCA_ and APV was weak. However, CBF_QCA_ in patients divided into three groups according to D_QCA_ showed a very strong correlation with APV.

### Significantly higher hMVRI for the RCA than for the LAD

In the present study, hMVRI was significantly higher for the RCA than for the LAD in both Groups A and B. Murai et al.^14^ reported that measurement at the RCA site, but not LAD or LCX, and hypertension were independent predictors of an increased IMR, according to the results for IMR in 131 arteries from 104 patients with intermediate obstructive coronary artery lesion. Similarly, Lee et al. ^15^ reported that 1096 patients with 1452 coronary arteries enrolled from 8 centers in 5 countries were analyzed by IMR, and predictors of high IMR included previous myocardial infarction, measurement at the RCA site, female sex, and obesity. IMR and hMVRI derive microvascular resistance from simultaneous distal coronary artery measurements of pressure and flow during hyperemia using intra-coronary guidewires. IMR estimates flow with thermodilution^3^, whereas hMVRI incorporates Doppler flow velocity^16^. Both indices have separately been shown to predict infarct size^17, 18^, microvascular obstruction^18^, regional wall motion^17^, and adverse left ventricular remodeling. Williams et al.^19^ reported that the correlation between independent invasive and noninvasive measurements of microvascular function was better with hMVRI than with IMR. However, neither of these invasive measures of microvascular resistance consider vessel size.

### Relevance of vessel diameter in evaluating CBF and HMRI

Unlike the results for APV, no significant difference in hMVRI_QCA_ was found for LAD, LCX, or RCA, respectively, when examined using CBF_QCA_. Furthermore, the correlation between APV and CBF_QCA_ was weak. Quantitative measurement of volumetric CBF (in milliliters per minute) during catheterization in humans is not currently possible. In evaluating CBF, a method that **accounts for** vessel size by QCA has been reported in addition to the semi-quantitative method using APV. Perera et al. evaluated angina and non-obstructive coronary artery disease by calculating epicardial diameter using QCA and calculating CBF for the purpose of evaluating endothelial-dependent microvascular function by acetylcholine^8^. In the present study, when vessel size was divided into three groups, a strong correlation was observed between CBFQCA and APV.

According to Doucette et al.^6^, APV showed a very strong linear correlation with coronary blood flow in small-sized straight tubes in vitro and in normal left circumflex coronary artery in dogs, but as diameter increased, slope decreased. Similarly, in the present study, the slope of the straight line between APV and CBF_QCA_ for small, medium, and large diameters in humans was 0.48, 0.30, and 0.21, respectively, decreasing as diameter increased. Moreover, we assessed associations between APV and CBF_QCA_ for coronary artery branches. Although the correlations between APV and CBF_QCA_ in the LAD and RCA were strong, that in the LCX was weak.

The LAD is the preferred target vessel in many studies, reflecting its myocardial mass and coronary arterial predominance, and much of the evidence for physiological assessments in INOCA has been obtained using the LAD.^7-9^ On the flip side, when the LAD cannot be used for technical reasons, use of the LCX is recommended, followed by the RCA. For measurements at the LCX, the orientation of the guide catheter makes injection of saline in the LCX direction difficult, and the guidewire, especially a Doppler and pressure sensor-equipped guidewire, is a stiff wire. This means that insertion into the LCX can be difficult due to the steep branch from the left main trunk to the LCX. Given this situation, we hesitate to perform measurements with anything other than the LAD because of the difficulty and potential for errors in measurement. In such situations, significant differences in microvascular coronary resistance may arise even among the three coronary branches. Measuring the index of microcirculatory resistance in multiple coronary territories may thus be useful, particularly in patients with typical angina in the absence of obstructive coronary artery disease. In the present study, similar to investigations using continuous coronary thermodilution^21^ or positron emission tomography (PET)^22-24^, interindividual variability in CBF_QCA_ and hMVRI_QCA_ values was observed. The main factor explaining the large ranges in flow and resistance values appears to be the dependence on myocardial mass. PET-derived flow and resistance express these measurements per unit of tissue mass (i.e., milliliters per minute per gram of tissue). Indeed, when adjusting coronary blood flow and microvascular resistance for mass, similar values were found for the three different myocardial territories within the same patients. Even so, even when adjusted for mass, inter-individual values for coronary blood flow and microvascular resistance show a considerable range.^23, 24^ Further confirming the hypothesis of natural variation in hyperemic myocardial perfusion, rather than measurement inaccuracies, Everaars et al.^24^ found hyperemic flow values in the range of approximately 50–450 ml/min in arteries. The large ranges of hyperemic flow values also found in the present study thus seem likely to correspond to a naturally occurring large variability of normal hyperemic values.

### Study Limitations

Several study limitations to this investigation should be acknowledged.

First, the present study of a relatively small population was performed at a single facility. Second, the papaverine intracoronary method is less well validated than the adenosine method and the results may not be the same as with adenosine.

Third, if the anatomy is highly tortuous in the coronary artery, adjusting the measurement site of the guidewire is technically difficult, resulting in inaccurate blood flow measurements. However, in the present study, all measurements were performed by operators with ample experience.

Fourth, the hMVRI is a reference standard measure of microvascular resistance and represents a useful parameter. The present study used two different methods to evaluate MR, including the traditional method (hMVRI) using APV and a method using vessel diameter (hMVRI_QCA_). We believe that hMVRI_QCA_ is the best composite clinical surrogate measure that can serve as a true reference standard.

Methods of calculating flow using APV are susceptible to several potential sources of inaccuracy.

APV depends on the assumptions that a time-averaged parabolic flow velocity profile occurs in which peak velocity is twice the mean velocity, that the peak of the flow velocity profile remains within the transducer’s sample volume throughout the cardiac cycle, and that the flow velocity profile remains unchanged with changes in flow rate. Normal proximal coronary arteries have been shown to have variable flow velocity profiles, the shapes of which may be almost perfectly parabolic, skewed toward the inner or outer wall, or trapezoidal^6,26-29^.

Near branches and in vessels that are tortuous or have intimal irregularities or stenoses, significant flow separation and deformation of the flow field is encountered^26,30^. In irregular, branched, or ectatic vessels, the relationship between peak and mean velocity is likely to be even more inconsistent. This may limit the usefulness of quantitative flow calculations based on APV, particularly in patients with diffuse coronary disease. Furthermore, unless accurate angiography is performed, inaccuracies in our estimations of vessel cross-sectional area may have occurred and may limit the reliability.

## CONCLUSIONS

Evaluation of coronary microvascular resistance in coronary branches of different diameters requires consideration of vessel diameter. While the LAD remains the preferred target vessel in hMVRI and IMR, when LAD cannot be used for technical reasons, RCA and then LCX are recommended by substituting coronary blood flow instead of APV.

## Data Availability

None

## Acknowledement

None

## Discussion

None

## Abbreviations List

hMVRI: hyperemic microvascular resistance index
APV: average peak velocity
CBF: coronary blood flow
QCA: quantitative coronary angiography
CMD: coronary microvascular dysfunction
INOCA: ischemia with no obstructive coronary arteries
MVA: microvascular angina
D_QCA_: vessel diameter analyzed by quantitative coronary angiography
CBF_QCA_: coronary blood flow by quantitative coronary angiography
hMVRI_QCA_: hyperemic microvascular resistance index by quantitative coronary angiography

